# Effectiveness and Cost-Effectiveness of TeleStroke Consultations to Support the Care of Stroke Patients Presenting to Regional Emergency Departments in Western Australia: An Economic Evaluation Case Study Protocol

**DOI:** 10.1101/2020.12.12.20248054

**Authors:** Christina Tsou, Suzanne Robinson, James Boyd, Shruthi Kamath, Justin Yeung, Stephanie Waters, Karen Gifford, Andrew Jamieson, Delia Hendrie

## Abstract

**Introduction:** The Western Australia Acute TeleStroke Programme commenced incrementally across regional Western Australia (WA) during 2016-2017. Since the introduction of the TeleStroke Programme, there has been monitoring of service outputs including regional patient access to tertiary stroke specialist advice and reperfusion treatment, however, the impact of consultation with a stroke specialist via telehealth (videoconferencing or telephone) on the effectiveness and cost-effectiveness of stroke care, and the drivers of cost-effectiveness has not been systematically evaluated.

**Methods and Analysis:** The aim of the case study is to examine the impact of consultation with a stroke specialist via telehealth on the effectiveness and cost-effectiveness of stroke and TIA care using a mixed methods approach. A categorical decision tree model will be constructed in collaboration with clinicians and programme managers. A before and after comparison using State-wide administrative datasets will be used to run the base model. If sample size and statistical power permits, the cases and comparators will be matched by stroke type and presence of CT scan at the initial site of presentation, age category and presenting hospital. The drivers of cost-effectiveness will be explored through stakeholder interviews. Data from the qualitative analysis will be cross-referenced with trends emerging from the quantitative dataset and used to guide the factors to be involved in sub-group and sensitivity analysis.

**Ethics and Dissemination:** Ethics approval for this case study has been granted from the WACHS Human Research and Ethics Committee (RGS3076). Reciprocal approval has been granted from Curtin University Human Research Ethics Office (HRE2019-0740). Findings will be disseminated publicly through conference presentation and peer-review publications. Interim findings will be released as internal reports to inform the service development.

**Strengths and Limitations of This Study:**

- Comparison of the impact of stroke specialist consultation via telehealth in regional Australia in supporting the management of different stroke subtypes
- The decision tree model will be constructed in collaboration with clinicians and programme administrators directly involved in the delivery of the TeleStroke Programme
- Use of local administrative data as model inputs enables the base model to reflect the reality of the regional WA Health service delivery
- Collaboration with WA Health stakeholders involved in TeleStroke Programme design and implementation to optimise utility of the case study to inform service development and expansion
- Conversion of the functional outcome modified Rankin Scale score (mRS) to quality adjusted life years (QALY) relies on national or international averages

## INTRODUCTION

Advances have been made in acute stroke management in recent years enabling ischaemic stroke patients to experience significant improved function and recovery post stroke^1^. The Western Australia Acute TeleStroke Programme commenced incrementally across regional Western Australia (WA) during 2016-2017. Since commencement, the number of WA rural patients receiving stroke reperfusion treatment that is mechanical thrombectomy (MT) and/or thrombolysis has increased across regional WA. Since the introduction of the TeleStroke Programme, there has been monitoring of service outputs including regional patient access to tertiary stroke specialist advice and reperfusion treatment, however, the impact of consultation with a stroke specialist via telehealth on the effectiveness and cost-effectiveness, and the drivers of cost-effectiveness has not been systematically studied.

The current body of knowledge on the effectiveness and cost-effectiveness of the telestroke services in rural and remote emergency departments (EDs) focuses on acute ischaemic stroke and the impact of telestroke on timely administration of thrombolysis and associated hospital resource use including inter-hospital transfers avoided. The studies have found telestroke to improve accuracy in decision making compared to telephone^2^ and to increase safe local administration of thrombolysis ^3^ with similar results to patients presenting directly to the tertiary stroke centres ^4^. However, moderate to severe ischaemic stroke patients treated with thrombolysis may potentially benefit from care at a tertiary stroke centre ^5^. Hence local thrombolysis administration does not completely mitigate the need for tertiary centre care. In addition, the increased accuracy in decision making also means that patients presenting to rural hospitals with more severe stroke (large vessel occlusion) who later transfer to tertiary hospitals can have equitable outcomes to patients who presented directly to the tertiary centre ^6^. Effectiveness studies typically focus on ischaemic stroke patients. The pattern and impact of telestroke access to haemorrhagic stroke, transient ischaemic attacks (TIAs) and stroke mimics from ED presentation to discharge is not well documented in the literature. ^7^

Medeiros De Bustos et al have recently observed that majority of telestroke studies concerns acute management^8^. There have been few socioeconomic studies to determine the true impact of telestroke in medical and economic terms^8^. There is also much to add to the literature on the association of patient outcome at discharge and in the longer term for each stroke subtypes to health systems cost.

Despite Whetten et al and Demaerschalk et al estimating telestroke access to be the dominant strategy compared to no access, the results are only valid when a long list of assumptions hold true ^7 9^. Cost-effectiveness analysis on rural and remote telestroke programmes to date have used effectiveness measures from previous clinical trials adjusted to a specific setting (e.g. ED), based on resource utilisation data obtained from published literature or publicly available data ^7 9-11^. These studies make assumptions or estimates from multiple data sources or rely on clinical expert opinions on activity variables such as probability of transfer to tertiary centres, discharge destination, and key resource utilisation variables such as cost of telestroke equipments^10 11^. These economic models have not taken into account haemorrhagic stroke or TIA patients who may benefit from telestroke ^10^, and did not consider secondary benefits of acute telestroke access on resource utilisation required within subacute episode of care following stroke and TIA^11^.

This study uses the TeleStroke Programme in Western Australia as a case study to examine the effectiveness and cost-effectiveness of telehealth to support the care of patients presenting to regional emergency departments in Western Australia. The primary interest in this research is twofold. Firstly, to understand the effectiveness and cost-effectiveness of the TeleStroke based on its current operation. Secondly, to gain insight into the drivers of cost-effectiveness of the service to inform future expansion of the TeleStroke Programme. The following research questions will be addressed:

1. Is seeing a stroke specialist via telehealth more effective and cost-effective than no stroke specialist involvement for patients presenting to rural and remote hospitals?
2. What are the drivers of cost-effectiveness in the above scenarios?
3. What factors other than access to stroke specialists via telehealth affect the clinical outcomes for patients presenting to a WACHS site with and without access to computed tomography (CT)?
4. What is the acceptability of stroke specialist consultation via telehealth to health professionals and health consumers and how do the enablers and barriers to its implementation help to explain the findings from the cost-effectiveness analysis?

## METHODS AND ANALYSIS

A mixed methods methodology will be used to allow incorporation of health care complexities into a systematic understanding of the factors that influence health service provision ^12^. A mixed methods approach is frequently used in health services research to allow quantitative data from survey and administrative records to combine with contextual information from qualitative data collections to support a better-informed decision about health service delivery. Guided by the model for assessment of telehealth (MAST)^13^, this case study will use a sequential explanatory design ^14^, with a combination of the methods described below. That is, the case study will commence with an economic evaluation of the TeleStroke Programme (quantitative phase), then conduct an analysis of its implementation which will be guided by a modified consolidated framework for implementation research (mCFIR) (qualitative phase), followed by the combined interpretation of the findings from the two phases of analysis to understand the drivers of cost-effectiveness.

The WA acute TeleStroke Programme industry stakeholders have been engaged in the design of the economic evaluation to incorporate current process of care into the case study to ensure the outcomes measured in this study are meaningful to future development of the programme. A quantitative assessment will be conducted on the effectiveness of stroke specialist consultation via telehealth on alignment to clinical protocols to achieve desirable clinical outcome, improved functional outcome for patients, and the impact of consultation with TeleStroke specialist on the diagnosis of stroke. Descriptive output from this analysis will also provide model input into the economic model.

A decision tree analytic model will be built to inform the calculation of the incremental cost-effectiveness ratio (ICER) for the three types of stroke presentations: haemorrhagic stroke, acute ischaemic infarction (AIS) and TIA when a TeleStroke specialist consultation has taken place. Complementing this information will be a novel qualitative data collection from relevant clinicians, health professionals, and policy makers to understand factors associated with implementation of TeleStroke Programme across WACHS facilities. In the explanatory phase, knowledge generated from the various parts of the research will be triangulated to help interpret the results and the effectiveness and cost-effectiveness analysis. This process will also contribute to improving the process of service delivery. Figure 1 presents a schematic diagram of the study methodology for the WA Acute TeleStroke case study.

**Figure 1.**
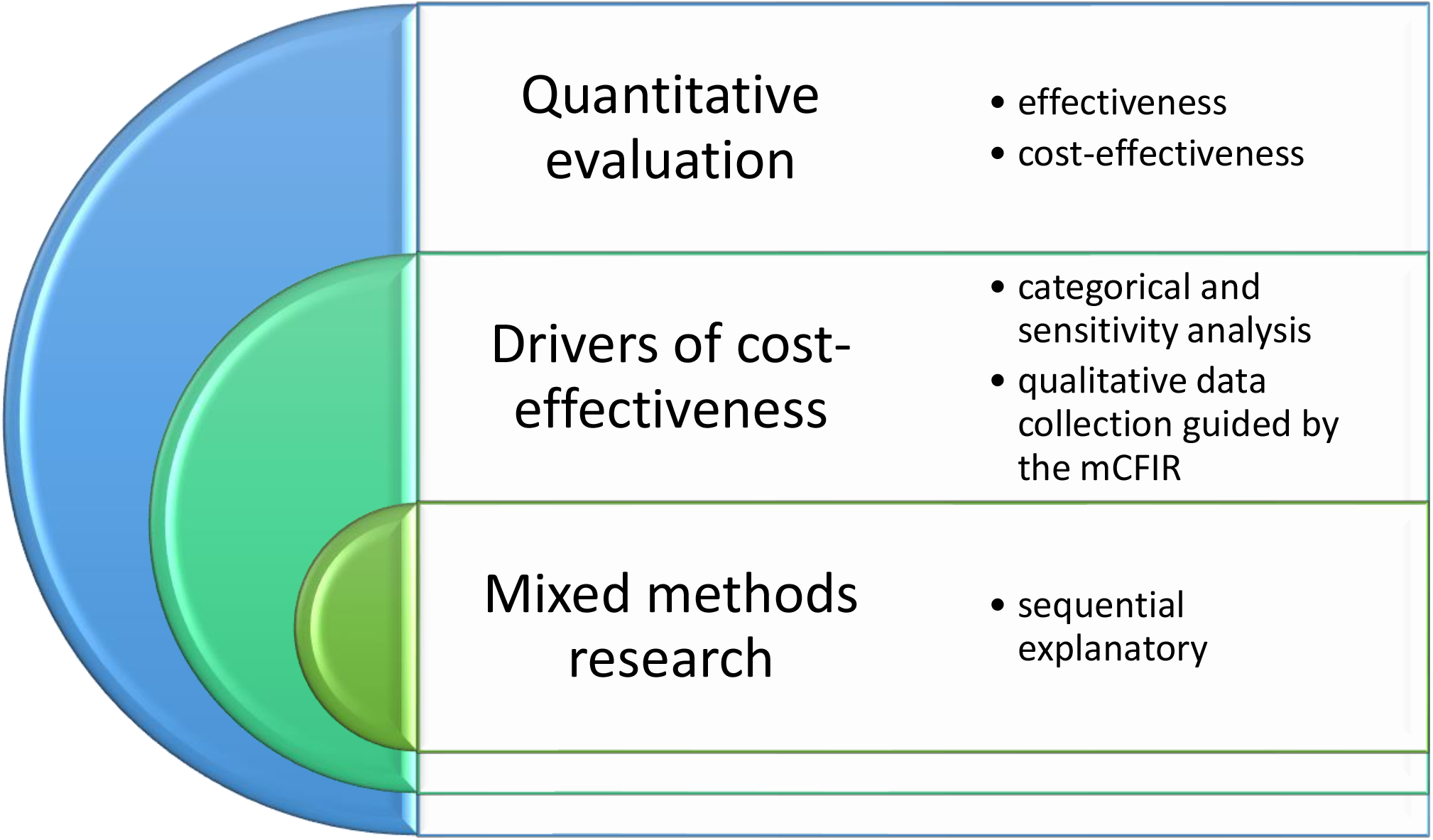
WA Country Health Service Acute TeleStroke Case Study Methodology.

### Case Study Settings

Medical emergencies in rural and remote Western Australia typically present to one of the WACHS facilities covering 2.5 million square kilometres. There are 12 sites across WACHS with computed tomography capabilities (CT sites) and 108 non-CT sites including 15 District Health Services, 49 small hospitals, 24 nursing posts / health centres, and 20 Aboriginal Community Clinics. Three geographically defined regional-metropolitan stroke pathways have been endorsed in WA linking WACHS facilities with partnered metropolitan acute stroke units. All WACHS emergency medical services are evaluated against the Emergency Care Capability Framework (ECCF) as the minimum requirements for role delineation capability and the minimum requirements to support the service.

### Administrative Data Collections

Six of the 12 CT sites across WACHS currently upload data to the WA Acute Stroke Data Collection (WAASDC). In this exploratory economic analysis, only sites contributing data to the WAASDC will be included. Learnings from this analysis will inform future economic evaluation designs across all WACHS sites. The WAASDC is a local file audit of stroke related indicators including whether a TeleStroke specialist was consulted. Other process-related indicators are available from the WACHS emergency department data collection (EDDC) and for admitted patients from the hospital morbidity data collection (HMDC), which are both routinely collected administrative datasets. By linking the three data sets, patients’ journey between WACHS hospitals will be studied.

To understand the patients journey to tertiary/quaternary care at other large or metropolitan sites, linkage to tertiary centre ED and admitted datasets will be done using patient identifier and date and time of discharge from WACHS hospital. The patient unique identifier will also be used to extract time and date of death from the State’s mortality data collection.

### Stroke case identification in the clinical process – an ICD-10 analysis

All stroke cases will be followed from the time of presentation to ED to the point of discharge from hospital. In-hospital stroke of admitted patients or admitted patients by-passing the ED will also be included in this analysis.

Stroke or TIA cases from the HMDC will be traced back to the EDDC to identify the principal diagnosis at ED. A separate stroke care audit was conducted on patients with established stroke or TIA diagnosis in the WAASDC. Patients identified as stroke or TIA cases in the EDDC but who were either discharged home or transferred to a tertiary centre for further treatment will be included as stroke cases. Those receiving consultation by a TeleStroke specialist after 1 January 2017 will be identified as cases and those presenting before 1^st^ of January 2016 are comparators. A 12 months window periods is to allow for data stabilisation. Descriptive analysis will then be performed on the cases and comparator subsets to provide model inputs to the cost-effectiveness analysis.

A subset of patients who have a stroke or TIA diagnosis recorded in the HMDC with a non-stroke/TIA diagnosis in EDDC will be extracted for analysis with presenting symptoms and whether consultations with TeleStroke specialists took place. This will give an understanding of the impact of TeleStroke on the diagnosis of Stroke in stroke patients’ hospital journey. The proportion of each type of change in diagnosis from EDDC to HMDC will be calculated and compared between the cases and comparator groups. This subset of patients will also be included as stroke cases for this analysis.

### Cost-effectiveness Analysis

Figure 2 summarises the components of the WA Acute TeleStroke Case Study. This study will use a decision tree analytic model to examine the incremental cost-effectiveness comparing the acute stroke care supported by TeleStroke consultations to acute stroke care prior to the implementation of the Acute TeleStroke Programme. The incremental cost-effectiveness ratio (ICER) will be calculated to determine the cost of increasing mRS by one unit, and another set of ICERs for cost per quality adjusted life year (QALY). Sub-group and sensitivity analysis will be performed to understand the drivers of cost-effectiveness and interpreted together with the qualitative data collection in the triangulation phase.

**Figure 2.**
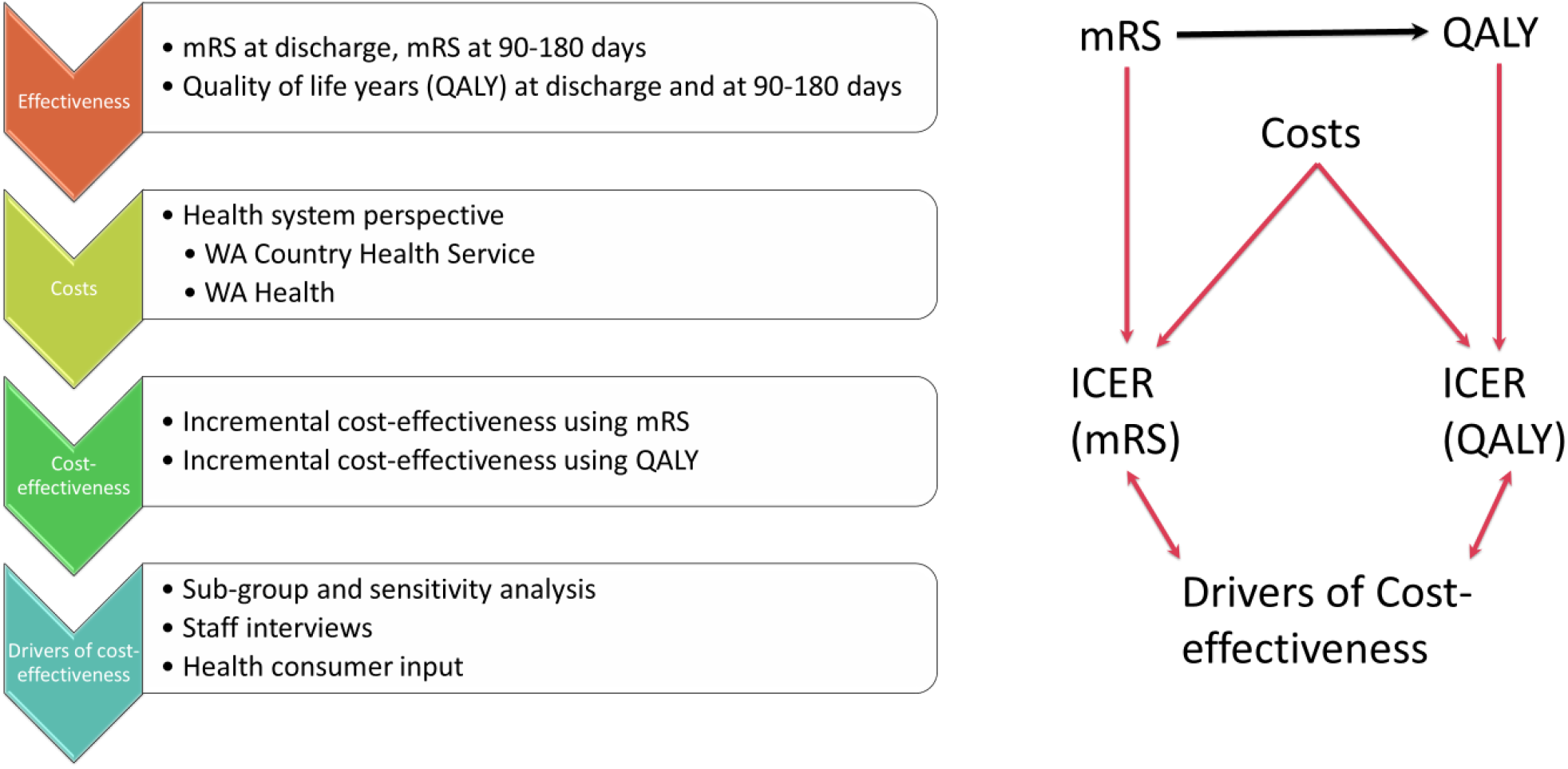
Components of the WA Acute TeleStroke Case Study.

#### Time horizon

There are three critical point in time for outcome determination in this study. Firstly, pre-intervention mRS to capture the functional status before thrombolysis or mechanical thrombectomy (MT). Secondly, at discharge, and finally at 90 to 180 days post discharge. The overall effectiveness of the stroke specialist telehealth consultations is determined by examining the change in functional status between pre-intervention and 90 to 180 days post discharge. The functional status at discharge is significant in examining whether the patient returned to the same functional state prior to the index stroke by comparing with premorbid mRS.

The WA Acute stroke pathways aligning the seven WACHS regions to tertiary hospitals commenced on the 1^st^ of January 2016. Patients presenting to WACHS ED from this point onwards can receive consultation from TeleStroke specialists. To allow a 12-month time window for data stabilisation, TeleStroke case selection will commence 12 months after the relevant site commence uploading to the WAASDC. The comparator selection will be from December 2015 to the earliest available records for the ED and HMDC data collections. This time horizon will be related to both the cost and effectiveness data collection.

#### Study perspectives

The analysis in this study will take the WACHS and WA public health systems perspectives by combining Country WA and metropolitan public hospital service utilisations, including TeleStroke Services, stroke unit care and neurological interventions.

#### The intervention and comparator

The study intervention in this study is stroke specialist consultation via telehealth. This is one component of the overall TeleStroke Programme. Given that TeleStroke consultations have been embedded within clinical workflows and processes, a contemporaneous non-Telestroke comparator group within hospital sites in this study would be subjected to selection bias with more TeleStroke use in higher acuity presentations. This, in addition to the vast geographical and community variations across WACHS regions, makes a contemporaneous comparator for the case study sites not feasible. Therefore, a pre and post study design is appropriate.

#### Index presentation and stroke case identification

Index presentations will be identified using the principal discharge diagnosis (ICD-10) from the WACHS ED and all diagnosis fields from the admitted data collections. Table 1 shortlists the ICD codes to be used to identify stroke presentations. Activity level model input to reflect care pathway will be collected from multiple data sources including the EDDC and HMDC and the WAASDC. Model inputs from the local data collections will be supplemented by the probability from the Australian Stroke Clinical Registry (AuSCR) or international averages published in peer reviewed literature to complete model inputs.

**Table 1.** Stroke and Transient Ischaemic Attack ICD-10-AM Sub-Classifications.

| Disease | Sub-category<br>(ICD-10-AM sub-classifications) | ICD-10-AM |
| --- | --- | --- |
| Stroke | Intracerebral haemorrhage | I61.0 – I61.6, I61.8 – I61.9 |
|  | Other non-traumatic intracranial haemorrhage | I62.9 |
|  | Cerebral infarction | I63.0 – I63.6, I63.8 – I63.9 |
|  | Stroke, not specified as haemorrhage or infarction | I64 |
| TIA | Transient cerebral ischaemic attacks, unspecified | G45.9 |

#### Decision model development

A categorical decision tree model will be developed in collaboration with WACHS clinicians and programme administrators capturing probabilities and decisions from WACHS ED presentation to the point of discharge from hospital. The modified Rankin Scale (mRS) at discharge will be the final outcome in the decision tree model. Time critical clinical outcomes (see outcome determination section below) will be used in a sensitivity analysis in the assessment of drivers of cost-effectiveness. Transition probabilities from presentation to a WACHS ED to discharge will come from the WACHS administrative datasets. The decision tree will be constructed by asking the following questions:

- Did a TeleStroke consult take place?
- Was there a bleed into the brain? (haemorrhagic vs non-haemorrhagic stroke)
- Was a major cerebral blood vessel blocked i.e. large vessel occlusion?
- Was the blood clot removed? (mechanical thrombectomy (MT) v no MT)
- Was treatment given to break up the blood clots? (thrombolysis v no thrombolysis)
- Did transfer to a tertiary centre take place? (transferred v admitted locally or managed locally)
- Were there complications associated with the treatment? (Intracerebral Haemorrhage (ICH) v no ICH)
- What was the functional status of the patient upon discharge and 90-180 days post discharge? (Modified Rankin Scale (mRS) cores 0-2 v 3-5 v 6)

#### Identification, measurement and valuation of resource use

Cost model calculations will be based on WA Health service and resource utilisation relating to the initial hospital presentation for stroke or TIA from presentation to hospital discharge, and from hospital discharge to 90-180-day post hospital discharge. Table 2 summarises the cost items.

**Table 2.** Cost Items for the WACHS Acute TeleStroke Case Study.

| Perspectives | Health Systems Perspective |  |
| --- | --- | --- |
|  | WACHS | WA Health |
| **Capital costs** | IT equipment | IT equipment |
| **Programme set up cost** | WACHS acute stroke programme costs | WA Stroke Services Project (WASSP) costs |
| **Direct health care costs** | ED service delivery cost<br>Hospital admission and rehabilitation cost<br>(include all direct patient care plus overheads comprising hospital, region and central office) | ED service delivery cost<br>Hospitalisation admission and rehabilitation cost<br>(include all direct patient care plus overheads) |
| **Direct non-health care costs (overhead costs)** | Overheads comprising hospital, region and central office. | Hospital overheads. |

##### Measuring Resource Use

Service cost will be collected from the WA Health costing system. The resource use will be built around individual patient encounters for the WACHS activity-based sites, these are the regional resource centres (RRC) and the district health services (DHS). All CT sites are either situated within a RRC or DHS therefore it is possible to build cost around individual patient encounters. Using the encounter identifier will allow integration of component direct health care costs supporting the episode of service delivery, and can potentially identify the breakdown of costs associated with the encounter including inpatient admissions, theatre and diagnostics services.

##### Valuing Resource Use

Unit cost data is available up to the 2018-19 financial year and all monetary values will be expressed in Australian dollars (AU$). All costs will be adjusted to mid-2018 value.

TeleStroke Programme costs will be evenly spread over the years since commencement of the WA Acute TeleStroke Programme. These include funding for tertiary specialists, project support and data collection through the Western Australia Stroke Services Project (WASSP). It will not be meaningful to apply an annual discount for programme costs as only two years of programme cost will be collected. ICT equipment costs will be depreciated on a straight line over their estimated useful lives.

#### Outcome Determination

##### Time Sensitive Measures

Stroke care is time sensitive, hence several time sensitive measures will be used to reflect the effectiveness of stroke protocol implementation with potential impact on patient outcome. Time sensitive measures showing strong association to functional outcomes for stroke patients will be identified as time-critical clinical effectiveness measures and used as a surrogate clinical outcome measure.

Door to imaging time indicates adherence to stroke care protocol, and door to first medical or nursing review, and door to TeleStroke consultation can impact patient outcome; while the length of hospital stay can reflect patient outcome and also goes directly to resource use. Onset to imaging time and onset to first medical or nursing review are the more clinically significant indicators; however, time of onset may not be accurately determined and it is expected that the time from onset analysis will be performed on a smaller subset of samples. Timely administration of thrombolysis to patients with ischaemic stroke is another significant factor impacting on patient outcome. This will be incorporated into the analysis if there is sufficient sample size for effectiveness analysis.

Clinical Interventions in haemorrhagic stroke are control of blood pressure and hemicraniectomy. Hemicraniectomy in WA will require transfer to a tertiary centre. Shorter door to tertiary centre transfer will be a meaningful time-critical clinical effectiveness measure. This will need to be interpreted in tandem with [regional ED] door to [tertiary centre] hemicraniectomy time to incorporate variations in transfer time (time transfer out of regional hospitals to arrival time at tertiary centre), and tertiary centre care.

##### Functional Outcome Measure

This study will use the modified Rankin Scale (mRS) score as the main functional outcome measure. A mRS score of 1-2 is considered a good functional outcome from stroke ^5 9^, mRS score of 6 is death, while mRS score of 3-5 indicates mild to severe disability / dependency requiring provision of carer support. This indicates costs occurring outside of hospital system, including inability to be employed and loss of productivity to society. Table 3 provides descriptions for the mRS scores.

**Table 3.** Modified Ranking Score (mRS) Descriptions.

| Score | Description |
| --- | --- |
| 0 | No symptoms at all |
| 1 | No significant disability despite symptoms; able to carry out all usual duties and activities |
| 2 | Slight disability; unable to carry out all previous activities, but able to look after own affairs without assistance |
| 3 | Moderate disability; require some help, but able to walk without assistance |
| 4 | Moderate severe disability; unable to walk without assistance and unable to attend to own bodily needs without assistance |
| 5 | Severe disability; bedridden, incontinent and require constant nursing care and attention |
| 6 | Dead |

The mRS score at discharge for patients presenting to WACHS ED prior to January 2020 will be retrospectively determined from discharge summary audits independently conducted by the WACHS Rural TeleStroke Neurology Fellow and WACHS TeleStroke Programme Manager and WACHS TeleStroke Programme Research Support Officer, who will discuss any discrepancies in scores to reach a consensus score. Similarly, the 90-180 day mRS for patients presenting to WACHS ED prior to January 2020 will be determined from subsequent public hospital utilisation audits.

The discharge destination rule in Nelson ^10^ will not be followed in this case study because anecdotal observation of WACHS patients indicate that patients who are discharged to residential aged care facility (RACF) are more likely to stay the same or deteriorate, while patient discharge home may improve with further rehabilitation. Nelson^10^ applies the following rules to determine the 90-180 day mRS score for patients who have not been readmitted to the public hospital system post discharge from index episode. The initial mRS score was assumed to improve by 1 point at 90 days for patients who were discharged from hospital to rehabilitation in the community. The initial mRS score was assumed to not change in patients who were discharged either home or to RACF ^10^. Across WACHS, if a patient is discharged to rehabilitation, majority in outpatient services, they may have improvement in mRS at 90-180 days. If the rehabilitation is provided by WA Health, mRS may be extracted from the patient’s medical record. For patients who have no further health system intervention post discharge, this study assumes that their mRS remains unchanged; however, there is also the possibility that they access non-WA Health rehabilitation and mRS improves. Deterioration may be marked by subsequent WA Health hospital admission.

##### Quality Adjusted Life Years (QALY) Estimation

This study will use the 90-180 day mRS score to estimate quality adjusted life years (QALY) gained for the above TeleStroke scenarios ^7 9 10^. QALY will be calculated using years of life remaining for each mRS at 6 months multiplied by utility weight sourced from CEA registry and their resulting sources^7 15-17^. The years of life remaining will be sourced from WACHS administrative data collections supplemented by national averages from AuSCR or previous studies ^18^. Utility weights will be measured on a 0 to 1 scale where 1 represents a state of perfect health and 0 represents death.

#### Estimated Sample Size

The Glick sample size formula^19^ has been used to estimate the sample size required for each of the TeleStroke and non-TeleStroke groups for this study to have 95% confidence with 80% study power. It is assumed that both groups will have equivalent standard deviation for cost and QALY, and equivalent sample sizes. The expected variations between the TeleStroke and non-TeleStroke groups have been sourced from previous cost-effectiveness analysis in the rural and remote context by Nelson et al^10^ and Whetten et al^7^.

Edney et al in 2017 calculated a reference willingness to pay of AU$28,033 per QALY gained and suggested that new technologies with ICER above AU$40,000 per QALY gained was recommenced for public funding^20^. Assuming the WA sample has equivalent variations in average resource use and QALY gained between the TeleStroke and non-TeleStroke groups, 607 individuals presenting with acute stroke episodes is required for each of the TeleStroke and non-TeleStroke groups if the public sector’s willingness to pay is AU$28,033 per QALY gain. This estimate of sample size would be 482 individuals if this willingness to pay is AU$40,000.

#### Analysis

##### Base case analysis and regression

Baseline characteristics and clinical and functional outcomes of the patients in the before TeleStroke and after TeleStroke groups will be summarised by stroke subtypes. Differences in resource use and costs between the two groups will be tested using two-sample t-test (or non-parametric equivalents) and Chi square tests for continuous and categorical variables, respectively. The mean costs of resource use in each care pathways in the decision tree and the difference in costs between the two groups will be calculated with 95% confidence interval.

##### Incremental cost-effectiveness

The primary comparison is between the TeleStroke and non-Telestroke groups and an overall incremental cost-effectiveness ratio (ICER) will be calculated using the formulae below at hospital discharge and at 90-180 day.

The first set of ICER will be calculated using the mRS as the effectiveness measure using the following:

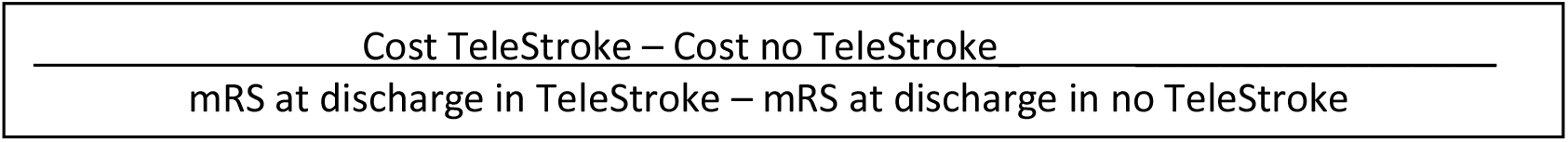

The second set of ICER will be calculated using QALY as the main outcome measure using the following:

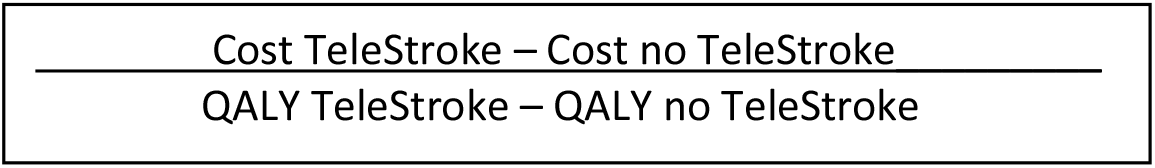

Subject to sufficient statistical power, a series of sub-group analyses will be performed to assess whether the cost and effectiveness association is dependent on a third variable. The initial list of sub-analyses is as follows and the final list will be guided by findings from the qualitative phase of the study:

1. Gender
2. Establishment
  a. Busselton Health Campus
  b. Bunbury Health Campus
  c. Albany Health Campus
  d. Geraldton Hospital
  e. Kalgoorlie Health Campus
  f. Northam Hospital
3. Recurrence: stroke recurrence categories
  a. First ever stroke
  b. Recurrent stroke <255 days (Western Australian median time to first stroke recurrence from first-ever stroke) ^21^
  c. Recurrent stroke >255 days
4. Rehabilitation post index presentation
5. Haemorrhagic stroke patients who underwent hemicraniectomy
6. Changes in Emergency Care Capability Framework (ECCF) domain achievement

The model results will provide estimates for the ICER considering health outcome and costs at discharge and at 90-180 days post discharge. The results of the economic evaluation including the sub-group and sensitivity analyses will be presented in terms of point estimates, cost-effectiveness planes and cost-effectiveness acceptability curves.

The stroke sub-types will be compared on the basis of incremental net monetary benefits. The net monetary benefit of the stroke specialist telehealth consultations with or without CT scans will be calculated as: the mean QALY multiplied by the acceptable threshold values for the QALY (add reference), minus the mean cost of the TeleStroke Programme and CT scans. The threshold value should be interpreted as the monetary value of a QALY.

##### Sensitivity Analysis

Sensitivity analysis will be performed to identify changes to ICER under a range of conditions. One-way analysis will be performed initially on varying the following variables: changes in TeleStroke consultation rates, changes in door to image time, changes in thrombolysis and MT numbers, changes in rate of transfer to CT sites, and changes in other time-sensitive outcomes listed above. The variables for inclusion in the multi-variate sensitivity analysis will be decided based on findings from the semi-structured interviews and preliminary quantitative analysis of the administrative datasets.

### Implementation Factors to Inform Future Expansion of Acute TeleStroke Programme Delivery in Western Australia

A qualitative data collection is incorporated into the study design to provide a robust evidence base to support the interpretation of findings from the effectiveness and cost-effectiveness studies described above. Guided by a modified consolidated framework of implementation research (mCFIR), this data collection is expected to provide a subsidiary evidence base informing the future development and implementation of the WA Acute TeleStroke Programme across WACHS.

#### The Modified Consolidated Framework for Implementation Research (mCFIR)

The Consolidated Framework for Implementation Research (CFIR) ^22 23^ has been chosen as the underpinning interpretative framework to guide the qualitative study (including design of interview protocol) and the final interpretation of the drivers of cost-effectiveness to adequately capture the complex nature of rural and remote health service delivery. As the focus of this research is on factors enabling and barring the successful integration of telehealth into clinical service implementation, the Finch, Mair et al theoretical constructs of normalization process theory (NPT)^24^ will add the system integration dimension and theoretical content. Key concepts in the MAST domains of preceding consideration, safety, patient perspectives, organisational, socio-cultural, ethical and legal aspects^13^ will also be mapped to the above ensuring coherence with the previous experience in telehealth evaluation. The resultant modified Consolidated Framework for Implementation Research (mCFIR) constructs will provide the initial categories in the qualitative analysis, and will be iteratively applied throughout the interpretive phase of the research.

#### Construction of interview guide

An interview guide has been developed based on the initial evaluation of the WA Acute TeleStroke programme implementation and impact within WACHS. Different sets of questions have been developed for the staff in the WACHS central office involved in the WACHS wide planning and implementation of the programme, programme sites with and without established pathways for stroke care. The draft interview protocol was reviewed by three stroke coordinators and senior managers within the WACHS Innovation and Development directorate.

#### Participant recruitment and the interview process

The aim of the qualitative component of this study is to explore and explain the factors impacting implementation of clinical telehealth strategies in rural and remote WA and the extent of their integration into routine clinical service delivery. The initial WACHS staff participants (seed participants) will be clinicians, programme administrators who have been involved in the commencement and implementation of the WA Acute TeleStroke Programme. The seed participants include senior leaders and managers, regional stroke coordinators, the neurology registrar. A snowball sampling technique will be used to identify and recruit further clinicians and health consumers to participate the study.

The semi-structured interviews will be conversational and interview protocol is designed for use as a guide to ensure consistent coverage of topic areas. Instead of asking the exact questions to every participant, the prompts under the main questions will be used as check lists to ensure relevant themes are covered in the dialogues. Small focus groups of less than 3 staff may be used instead of one on one interview when required to meet the constraints of participants’ schedules.

The interviews will be audio-recorded and transcribed verbatim and managed in NVivo. While the open coding is performed, notes will be taken under the modified CFIR domain and construct headings. Axial coding will then be performed to make interconnections between the emerging themes within each CFIR construct. These relationship will be pulled into a relational model of causal conditions, central phenomenon, context, intervening conditions, action/interaction strategies and consequences ^25^. Selective coding will then be performed to connect the emerging themes to produce a discursive set of theoretical propositions on telehealth integration in clinical service delivery ^25^. The simultaneous data collection and analysis process will continue until data saturation is reached for all relevant constructs in the modified CFIR framework.

### Drivers of Cost-effectiveness and Generation of Deliberative Dialogue

This case study will use two methods to gain insights into the drivers of cost-effectiveness. It will be an iterative process between the sub-group and sensitivity analyses part of the economic analysis, and the qualitative exploration. These two processes will commence as separate and concurrent parts in the case study. A constant comparison of early findings will take place where key themes emerging from the dialogues and interviews with key industry stakeholders, and health consumers is expected to offer explanation to the findings from the sub-group and sensitivity analyses. Any new categories or factors emerging from this constant comparison of findings will feed into the sub-group and sensitivity analyses as new sub-categories or factors for consideration in assessing the level of confidence of the ICERs calculated (whether the cost-effectiveness finding is robust when certain factors vary individually or in combination).

The understanding of the drivers of cost-effectiveness will include tacit knowledge of clinician, practitioners and consumers through deliberative dialogues generated during the final explanatory phase i.e. interpretive phase of the research. The deliberative dialogues generated in the interview and dissemination of early findings is likely to create an evaluative practice supporting the sharing of best practice and informing future rural and remote telehealth planning and policy in WA Health.

### Patient and Public Involvement

A consumer representative was involved in the conceptualisation of this protocol.

## Data Availability

As this is a protocol paper, data has not yet been collected.

## ETHICS AND DISSEMINATION

Ethics approval for this case study has been granted from the Western Australian Country Health Service Human Research and Ethics Committee (WACHS HREC Project Reference Number RGS3076). Reciprocal approval granted from Curtin University Human Research Ethics Office (approval number HRE2019-0740). Findings will be disseminated publicly through conference presentation and peer-review publications. Interim findings will be released as internal reports to inform the TeleStroke Programme development.

## AUTHOR’S CONTRIBUTIONS

Christina Tsou drafted and provided the initial design of study. She has also coordinated input from the authorship to bring the work to publication. Delia Hendrie and Suzanne Robinson provided input on health systems and health economic methodology. James Boyd provided input on statistics and data linkage. Andrew Jamieson provided high level clinical practice perspective in the rural and remote Australia. Justin Yeung brings regional emergency medicine and telehealth application perspective to this project. Shruthi Kamath contributed specialist neurology knowledge and provided significant input in the methodology related to clinical effectiveness measures. Stephanie Waters and Karen Gifford provided programme implementation perspective and made significant contribution to the design of the interview protocol. All authors have reviewed the final manuscript and agreed to submit for publication.

